# Targeted follicular fluid proteomics using reverse phase protein arrays (RPPA); a feasibility study

**DOI:** 10.64898/2026.02.02.26345389

**Authors:** Michael S. Bloom, Veronica G. Sanchez, Victor Y. Fujimoto, Makeda Tamrat, Jenna R. Krall, Virginia Espina

## Abstract

This small pilot feasibility study shows that reverse phase protein array (RPPA) technology is a useful tool for targeted proteomics analysis in human ovarian follicular fluid. RPPA supplements mass spectrometry approaches that are currently used by providing functional signal transduction data that drive cellular biology. Herein, we present the first report of using RPPA in follicular fluid to elucidate protein signaling pathways. The results show potential associations between follicular fluid proteins measured with RPPA and reproductive outcomes from *in vitro* fertilization, including oocyte maturity, oocyte fertilization, embryo quality, and pregnancy. This study provides evidence that RPPA is a feasible approach to be used in clinical studies of reproductive endpoints. However, a larger study of RPPA to identify diagnostic and prognostic follicular fluid protein biomarkers of infertility is needed.

## Introduction

More than 20% of married and cohabiting women without a previous birth, approximately 5 million US women, experienced infertility between 2015 and 2019 [1]. Infertility, defined as 12 months of unprotected intercourse without a pregnancy, has driven increased use of *in vitro* fertilization (IVF) [2]. In 2022, US clinics initiated over 206,304 IVF cycles with intended embryo transfers, resulting in 98,289 live births, yet more than 110,000 IVF cycles failed [3]. Many women require multiple IVF cycles, facing psychological distress [4,5], higher obstetrical risks [6,7], and potential long-term adverse health consequences [8–11], along with financial costs that often exceed $19,000 per cycle [12]. Transferring multiple embryos to improve the chances of a live birth also raises the risks of multiple gestations and adverse birth outcome [13–15]. With two-thirds of IVF cycles failing to produce a live birth, there is a critical need to identify biological targets to improve outcomes [3,16,17].

Proteomic profiling of follicular fluid (FF) collected and retained during IVF can offer direct measures of critical proteins that drive oocyte maturation and the subsequent developmental events that eventually lead to live birth. While great advances have been made in describing the FF proteome, results have been inconsistent and few clinically-actionable data have emerged [18]. Mass spectrometry (MS) methods have identified highly abundant FF proteins and peptides, but these have limited sensitivity for post-translationally modified proteins, and require extensive expertise and data processing, large sample volumes, and complementary multiple reaction monitoring with heavy isotope labeled peptides for quantification.

As a complementary approach to MS, reverse phase protein arrays (RPPA) offer a cost effective and multiplex strategy that has been extensively validated with biological fluids and tissues and is used in oncology clinical trials for developing precision tumor treatments [19–23]. However, RPPA has not been previously reported with FF. RPPA allows analysis of small sample volumes (e.g., nanograms), with the sensitivity (e.g., femtograms) and specificity of monoclonal antibodies, while quantifying proteins and protein modifications that govern their biological activity, such as phosphorylation, acetylation, or cleavage [22]. To assess the feasibility, we conducted an outcome-blinded pilot assessment of 21 FF proteins from critical biological pathways selected *a priori* in 6 FF specimens from women undergoing IVF.

## Methods

### Study Protocol

Up to 4 independent FF samples were collected from 56 women undergoing IVF and enrolled in the Study of Metals and Assisted Reproductive Technologies (SMART) in 2015-2017. The study protocol was described in detail in a previous publication [24]. Briefly, women underwent gonadotropin-induced ovarian stimulation with serial ultrasounds and estrogen measures. Human chorionic gonadotropin was administered approximately 2 weeks later, after follicles developed to ≥17mm diameter, and oocytes were retrieved after 34-36 hours by transvaginal fine needle aspiration. In each ovary, the largest follicle was aspirated; after evacuation, the needle was flushed with saline before sampling a second follicle, but the follicle itself was not flushed to preserve native analyte concentrations. After the oocyte was removed, each individual 3.5-5mL FF sample was centrifuged to pellet residual cells and debris and aliquoted the supernatant into 1.8mL cryovials, which were frozen at -80 °C. Any samples showing evidence of red blood were discarded [25].

Mature oocytes collected from ovarian follicles in metaphase-2 arrest (MII arrest) were fertilized with sperm using intracytoplasmic sperm injection or conventional insemination. Fertilization was confirmed after 16–20 hours by the appearance of 2 pronuclei (2PN). Embryo and blastocyst quality were categorized as “high quality” or “low quality’ based on a day 2 or 3 embryo examination of blastomere fragmentation, cleavage rate, and blastomere symmetry [26–30], or Gardner’s scale for day 5 blastocysts [31]. Fresh embryos or blastocysts were transferred and a positive serum hCG test 2 weeks later indicated pregnancy. All participants completed written informed consent and the study protocol was approved by the UCSF Committee on Human Research.

### Reverse Phase Protein Arrays (RPPA)

RPPA analysis was used to quantify 21 FF proteins, normalized to total FF protein concentration, in a single FF from 3 women with an IVF pregnancy and 3 without an IVF pregnancy, randomly selected from SMART participants. FF was printed in two-fold serial dilutions on a nitrocellulose-coated slide (Grace BioLabs, Oncyte Avid, Bend, Oregon, USA) that included calibrators and controls for rigorous, clinical diagnostic level analysis. Twenty-one FF proteins were selected *a priori* based on validated antibodies available in our Center from the endocannabinoid system, oxidative stress and inflammatory response, epigenetic markers, tryptophan metabolism, cell division and migration, hormone synthesis and function, DNA damage and repair, and vitamin D homeostasis pathways as described in **Table 1**.

**Table 1.** Proteins analyzed in human ovarian follicular fluid using RPPA.

| Protein | Abbreviation | Source |
| --- | --- | --- |
| Tripeptidyl peptidase 1 | TTP1 | Cell Signaling Technology |
| Nuclear factor-κB | NFKB | Cell Signaling Technology |
| P53 binding protein | P53BP1 | Cell Signaling Technology |
| Vitamin D binding protein | VitDBP | Abcam |
| Interleukin-1 $\beta$ | IL-1b | Cell Signaling Technology |
| Estrogen receptor $\beta$ | ERbeta | DSHB |
| Copper-Zinc superoxide dismutase | CuZnSOD | StressGen Biotechnologies |
| Cleaved caspase 3 (Asp175) | CICasp3 | Cell Signaling Technology |
| Prolactin receptor | ProR | Epitomics |
| Phosphorylated (Tyr 194) peroxiredoxin 1 | Prdx-1 | Cell Signaling Technology |
| Manganese superoxide dismutase | MnSOD | Assay Designs |
| Interleukin-6 | IL-6 | BioVision |
| Wingless integrated 5a/5b | Wnt5ab | Cell Signaling Technology |
| Vitamin D3 receptor | VitDR | Cell Signaling Technology |
| Peroxisome proliferator-activated receptor $\gamma$ | PPARgamma | Cell Signaling Technology |
| Phosphorylated insulin-like growth factor receptor $\beta$ (Tyr 1135/36) | IGF1R-beta | Cell Signaling Technology |
| Indoleamine-pyrrole 2,3-dioxygenase | IDO | Cell Signaling Technology |
| Cleaved histone deacetylase 3 | HDAC3 | Cell Signaling Technology |
| Bloom syndrome protein | BLM | Cell Signaling Technology |
| Glutamine dehydrogenase | GDH | Cell Signaling Technology |
| Diacylglycerol lipase $\alpha$ | DAGLipalpha | Cell Signaling Technology |

Abbreviations: RPPA, reverse phase protein arrays

Each array was probed with a validated, commercially available monoclonal or polyclonal antibody. Antibody validation is performed for each antibody and whenever a new lot number of antibody is received, following CAP/CLIA compliant standard operating procedures. There is a compendium of more than 400 validated antibodies in our Center. Each sample, control, and calibrator was printed in technical replicates on the arrays (required coefficient of variation < 15%) [32] (**Figure 1**). Bovine serum albumin was used as the total protein calibrator. Commercial cell lysates served as process controls. Calibrators, created from cell lines, spanned the limits of quantification for the proteins of interest. The calibrators may also be used to interpolate the relative intensity values of each spot on the array. The pixel intensity of each spot was quantified using a calibrated flatbed scanner (UMAX PowerLook, UMAX Technologies, Dallas, Texas, USA) for chromogenic detection or a laser scanner (Tecan Group Ltd., Zurich, Switzerland) for fluorescent detection.

**Figure 1.**
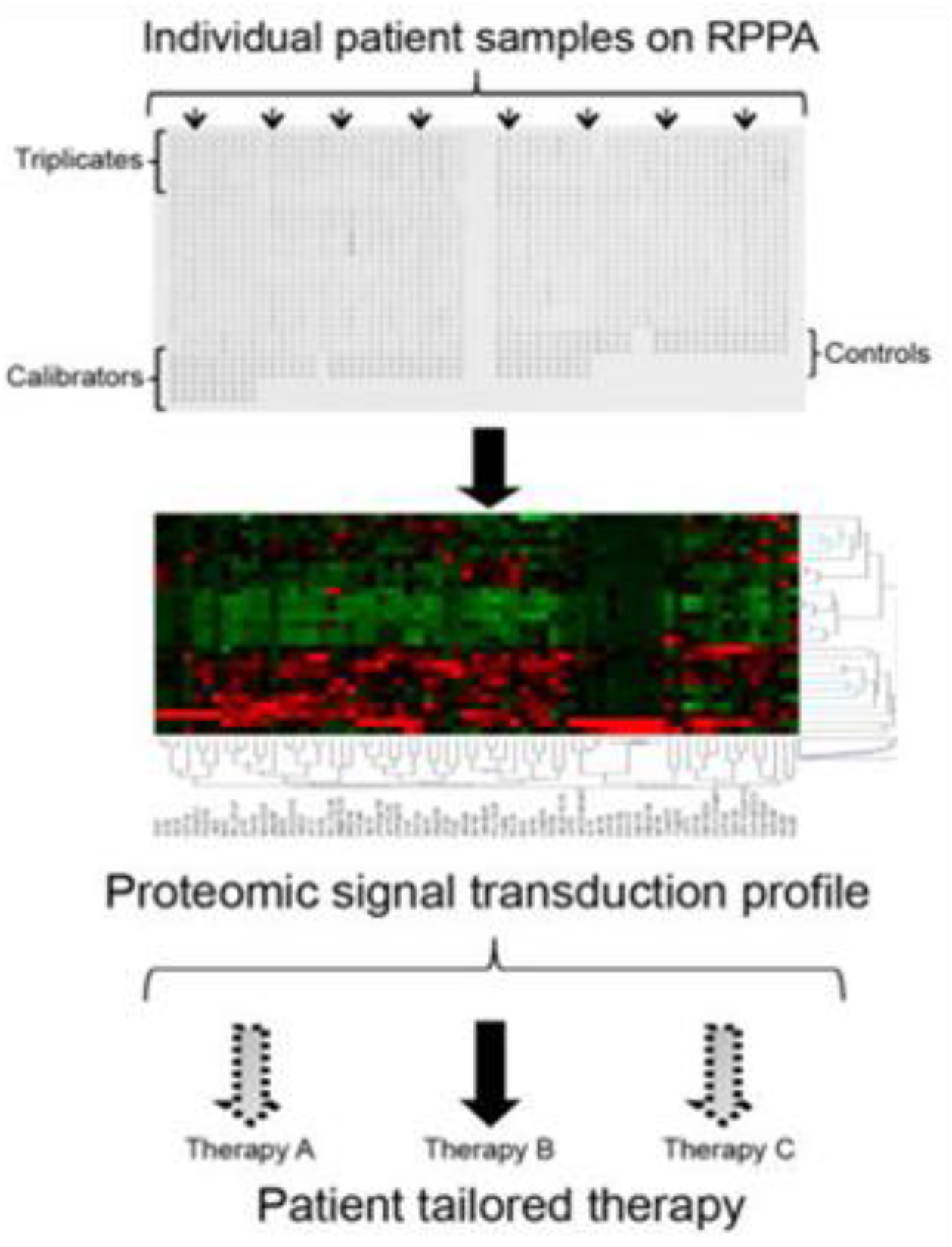
Reverse Phase Protein Arrays (RPPA) provide quantitative data for cell signaling kinases and their post-translational modified forms. Specimens, control, and calibrators are printed on nitrocellulose coated slides in replicate dilution curves. Each array is probed with a single, validated antibody and catalyzed signal amplification chemistries.

Signal intensities greater than 3 standard deviations above background were considered to have an adequate signal-to-noise ratio for the analysis [33]. RPPA spot intensity was determined using ImageQuant v.5.2 software (Cytiva, Marlborough, Massachusetts, USA). The local area background was subtracted from each spot and the data were normalized to total protein using an in-house VBA Macro (RPPA Analysis Suite) [33].

### Data Analysis

We estimated the associations between FF proteins, oocyte quality, and embryo quality outcomes using Spearman correlation coefficients and compared differences in mean FF protein concentrations between women with and without an IVF pregnancy. We defined statistical significance as P<0.10 for a 2-tailed test to accommodate the limited sample size and generate hypotheses for future confirmation.

## Results

Seventeen of 21 FF proteins were quantified using RPPA in our CAP/CLIA-accredited proteomics laboratory, including TTP1, NFKB, p53BP1, Vitamin D Binding Protein, IL-1b, Cleaved Caspase3, Prolactin Receptor, Prdx-1, Manganese Super Oxide Dismutase, IL-6, Wnt5ab, Vitamin D Receptor, PPARgamma, IGF1R-beta, IDO, BLM, and DAG Lipase alpha (abbreviations are defined in **Table 1**). FF glutamine dehydrogenase, HDAC3, CuZnSOD, and ERβ were not quantifiable.

As shown in **Figure 2**, we found a mixed pattern of moderate to strong pairwise Spearman correlations between FF proteins and intermediate IVF outcomes. Oocyte maturity (MII arrest) and fertilization (2PN) correlated negatively to TPP1 and P53BP1, but positively correlated to NFKB, VitDBP, IL-1b, ClCasp3, ProR, Prdx-1, MnSOD, IL-6, Wnt5ab, VitDR, PPARgamma, IGF1Rbeta, and IDO. In contrast, greater TPP1 and P53BP1 correlated to high cleavage stage embryo quality, but greater FF VitDBP and IGF1Rbeta were associated with low cleavage stage embryo quality. There were no statistically significant correlations with blastocyst quality, which may have been due to the limited number of high quality blastocysts in the sample (i.e., 6 high quality blastocysts of 42 total blastocysts).

**Figure 2.**
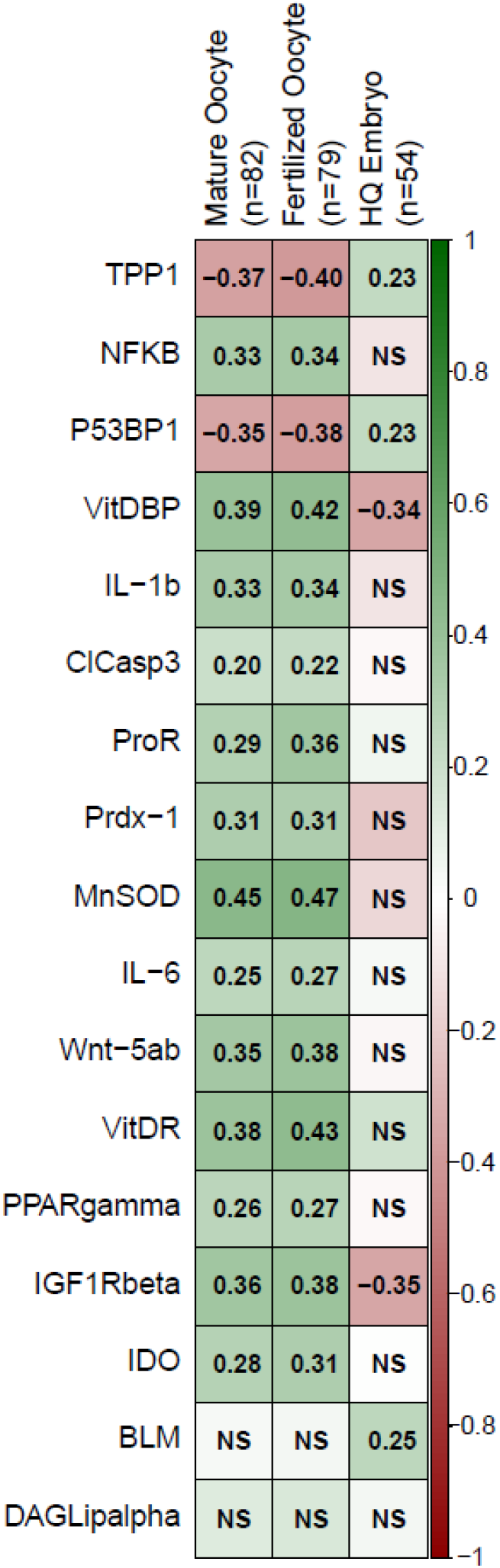
Spearman correlation coefficients between follicular fluid proteins measured using RPPA and intermediate IVF outcomes among women using IVF. Abbreviations: BLM, Bloom syndrome protein; ClCasp3, cleaved caspase 3; DAGLipalpha, diacylglycerol lipase α; HQ, high quality; IDO, indoleamine-pyrrole 2,3-dioxygenase; IGF1Rbeta, phosphorylated insulin-like growth factor receptor β; IL-1b, interleukin-1 β; IL-6, interleukin 6; IVF, *in vitro* fertilization; MnSOD, manganese superoxide dismutase; NFKB, nuclear factor kappa β; NS, not significant; P53BP1, P53 binding protein; PPARgamma, peroxisome proliferator-activated receptor γ; Prdx-1, peroxiredoxin 1; ProR, prolactin receptor; RPPA, reverse phase protein array; TPP1, Tripeptidyl peptidase 1; VitDBP, vitamin D binding protein; VitDR, vitamin D3 receptor; Wnt5ab, Wingless integrated 5a/5b NOTE: Colors in the boxes correspond to the Spearman correlation coefficients between individual follicular fluid proteins and the intermediate IVF outcomes (n for each test listed in each column), Darker green indicates a more positive correlation and darker red indicates a more negative correlation. Statistical significance was defined as P<0.10.

As shown in **Figure 3**, we found greater mean levels of NFKB (37.3%), P53BP1 (25.5%), VitDBP (29.8%), IL-1b (42.8%), ClCasp3 (35.6%), ProR (18.0%), Prdx-1 (15.6%), MnSOD (70.7%), IL-6 (43.0%), Wnt5ab (70.8%), VitDR (81.3%), IGF1R-beta (535.9%), and DAGLipalpha (44.2%) among non-pregnant than pregnant women. However, the sample size was too limited for formal hypothesis testing.

**Figure 3.**
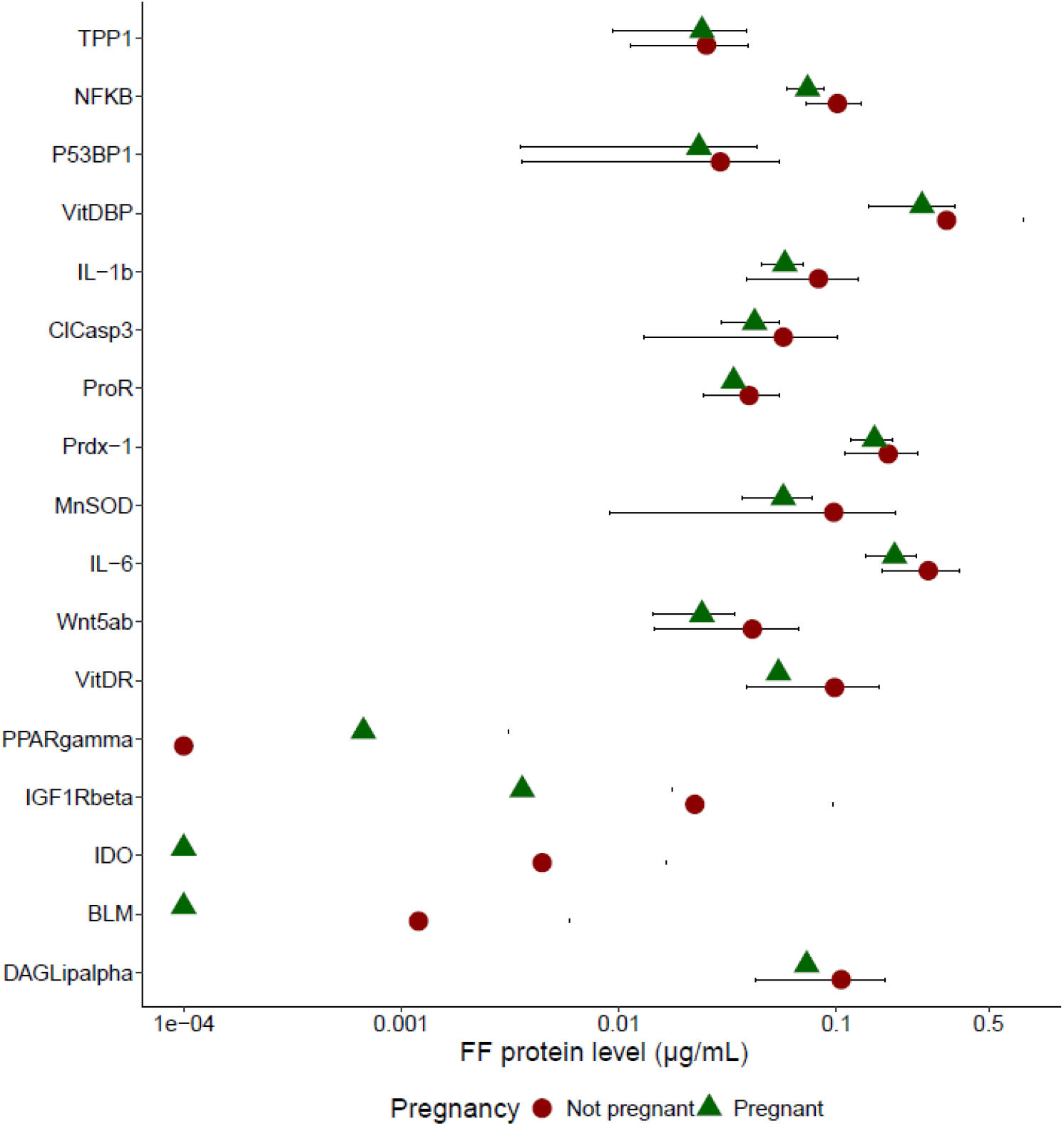
Mean (95% confidence interval) follicular fluid protein concentrations measured using RPPA, between women with (n=3) and without (n=3) an IVF pregnancy. Abbreviations: BLM, Bloom syndrome protein; ClCasp3, cleaved caspase 3; DAGLipalpha, diacylglycerol lipase α; HQ, high quality; IDO, indoleamine-pyrrole 2,3-dioxygenase; IGF1Rbeta, phosphorylated insulin-like growth factor receptor β; IL-1b, interleukin-1 β; IL-6, interleukin 6; IVF, *in vitro* fertilization; MnSOD, manganese superoxide dismutase; NFKB, nuclear factor kappa β; P53BP1, P53 binding protein; PPARgamma, peroxisome proliferator-activated receptor γ; Prdx-1, peroxiredoxin 1; ProR, prolactin receptor; RPPA, reverse phase protein array; TPP1, Tripeptidyl peptidase 1; VitDBP, vitamin D binding protein; VitDR, vitamin D3 receptor; Wnt5ab, Wingless integrated 5a/5b NOTE: Symbols correspond to mean follicular fluid protein concentrations (n=3 in each pregnancy group) and whiskers represent 95% confidence intervals. Symbols without whiskers represent uniformly measured values for a pregnancy group (i.e., BLM, IDO, PPARgamma), or measurement of n=1 or n=2 for a pregnancy group (i.e., BLM, IDO, IGF1Rbeta, PPARgamma, VitDR, VitDBP).

## Discussion

In this small feasibility study, we found different FF protein concentrations according to oocyte fertilization, embryo quality, and pregnancy outcomes using RPPA. Our results suggest that FF proteins in the endocannabinoid system (DAG-lipase α findings) [34–36], Wnt signaling pathway (Wnt5ab findings) [37,38], acute phase immune response (IL-6 and NFKB findings) [39,40], oxidative stress response (MnSOD and p53BP1 findings) [41,42], innate immune response (IL-1β and NFKB findings) [40,43], and vitamin D signaling pathway (VD3R and VDBP findings) [44–46] may be important determinants of IVF outcomes. However, these results are preliminary and a larger and more comprehensive analysis will be necessary for definitive results.

The potential for an oocyte to complete meiosis, fertilize normally, and support early embryo development, collectively referred to as oocyte maturation, depends on nuclear, epigenetic, and cytoplasmic processes that influence IVF outcomes [47,48]. Throughout development, oocytes are immersed in FF, which mediates nutrient exchange and biochemical communication with mural granulosa cells, cumulus cells, and the vascular compartment [49,50]. FF contains a plasma ultrafiltrate restricted by the blood–follicle barrier to molecules less than ~300 kDa, along with factors produced *in situ* by granulosa cells and the oocyte itself, creating a specialized microenvironment [51–53]. For example, 22 FF proteins differed between women with and without ovarian pathology in a recent study, while only 2 were detectable in matched serum [54]. Because FF is the fluid most proximate to the oocyte, it provides a unique window into the microfollicular environment and can reveal protein markers predictive of IVF outcomes that are diluted or undetectable in peripheral blood [49,55].

Proteomic profiling of FF collected during IVF provides insight into the protein systems that drive oocyte maturation and the downstream developmental events that lead to pregnancy and live birth [49,56,57]. Unlike genomic or transcriptomic assays, which may correlate only weakly with protein abundance or activation status, proteomic approaches directly characterize follicular phenotype in real time [58]. Over the past 15 years, FF proteome profiling has expanded substantially [17,56]. Investigators have increasingly identified and quantified proteins from major biological pathways in efforts to develop diagnostic and prognostic biomarkers. A synthesis of 617 FF proteins reported through 2014 highlighted acute phase inflammatory response, wound response, complement, coagulation, lipid metabolism, and cytoskeletal pathways as most frequently represented, with matrix metalloproteinases (MMPs), thrombin, and vitamin D/Retinoid X Receptor alpha emerging as central hubs [59]. Another study detected 742 proteins in pooled FF from 3 ovum donors using MS, primarily related to growth factor and receptor signaling, immunity, anti-apoptosis, MMP activity, and complement [60]. An MS-based biological pathway analysis similarly confirmed the prominence of complement and coagulation systems, with additional emphasis on innate immunity and angiogenesis [61].

More recent studies have expanded the FF proteome further. One MS analysis identified 2461 FF proteins, including 1108 detected for the first time, enriched in metabolic processes and biological regulation [62]. A SWATH (Sequential Window Acquisition of all Theoretical Mass Spectra)-MS study detected 2182 FF proteins, most lacking known metabolic functions, and others involved in coagulation, integrin signaling, gonadotropin-releasing hormone signaling, plasminogen activation, and Wnt signaling [63]. Despite substantial progress, findings across studies remain inconsistent and few clinically actionable biomarkers have emerged [18,49,56,59,64].

While MS-based approaches have identified highly abundant FF proteins and peptides, these methods require complex, specialized workflows for measuring low-abundance proteins and peptides in FF and their post-translational modifications [65]. The results of this feasibility study suggest that RPPA is a feasible complementary technology to quantify low-abundance ovarian FF proteins and their post-translationally modified forms that predict IVF outcomes, and may serve as diagnostic/prognostic indicators and targets for interventions. However, a larger validation study will be necessary to confirm that RPPA technology is a feasible approach to investigate FF proteins as potential clinical and prognostic indicators of IVF outcomes or targets for clinical intervention.

## Data Availability

The datasets generated and analyzed during the current study are not publicly available due confidentiality concerns, but are available from the corresponding author on reasonable request.

## Notes

### Competing Interest Statement

The authors have declared no competing interest.

### Funding Statement

This work was funded in part by the National Institute of Environmental Health Sciences (NIEHS), grant number 1R56ES023886-01 (MSB).

### Author Declarations

IRB of University of California San Francisco gave ethical approval for this work.

## References

[1] Nugent CN, Chandra A. Infertility and impaired fecundity in women and men in the United States, 2015-2019. Natl Health Stat Rep 2024:1–19.

[2] Peipert BJ, Adashi EY, Penzias A, et al. Global in vitro fertilization utilization: How does the United States compare? FS Rep 2023;4:326–7. 10.1016/j.xfre.2023.06.005.

[3] Center for Disease Control and Prevention. National ART Summary, 2022 2024. https://www.cdc.gov/art/php/national-summary/index.html x(accessed April 1, 2025).

[4] Braverman AM, Davoudian T, Levin IK, et al. Depression, anxiety, quality of life, and infertility: a global lens on the last decade of research. Fertil Steril 2024;121:379–83. 10.1016/j.fertnstert.2024.01.013.

[5] Peterson SK, Jennings Mayo-Wilson L, Spigel L, et al. Health care experiences of individuals accessing or undergoing in vitro fertilization (IVF) in the U.S.: a narrative review of qualitative studies. Front Reprod Health 2025;Volume 7-2025.

[6] Sullivan-Pyke CS, Senapati S, Mainigi MA, et al. In vitro fertilization and adverse obstetric and perinatal outcomes. Perinat Epidemiol 2017;41:345–53. 10.1053/j.semperi.2017.07.001.

[7] Bentov Y, Schenker J. IVF and pregnancy outcomes: the triumphs, challenges, and unanswered questions. J Ovarian Res 2025;18:228. 10.1186/s13048-025-01692-5.

[8] Nguyen HT, Hanevik HI, Fraser A, et al. Women’s risk of hypertension and cardiovascular disease subtypes by number of cycles of assisted reproductive technologies: a Norwegian registry-linkage study. Eur J Prev Cardiol 2025:zwaf719. 10.1093/eurjpc/zwaf719.

[9] Saso S, Barcroft JF, Kasaven LS, et al. An umbrella review of meta-analyses regarding the incidence of female-specific malignancies after fertility treatment. Fertil Steril 2025;123:506–19. 10.1016/j.fertnstert.2024.09.023.

[10] Liang Z, Wang D, Wang X. Cardiovascular safety of assisted reproductive technologies: what have we learned and what remains unknown? Eur Heart J 2025;46:1867–8. 10.1093/eurheartj/ehaf108.

[11] Pivato CA, Inversetti A, Condorelli G, et al. Cardiovascular safety of assisted reproductive technology: a meta-analysis. Eur Heart J 2025;46:687–98. 10.1093/eurheartj/ehae886.

[12] Peipert BJ, Montoya MN, Bedrick BS, et al. Impact of in vitro fertilization state mandates for third party insurance coverage in the United States: a review and critical assessment. Reprod Biol Endocrinol 2022;20:111. 10.1186/s12958-022-00984-5.

[13] Bergh C, Kamath MS, Wang R, et al. Strategies to reduce multiple pregnancies during medically assisted reproduction. Fertil Steril 2020;114:673–9. 10.1016/j.fertnstert.2020.07.022.

[14] Adamson GD, Norman RJ. Why are multiple pregnancy rates and single embryo transfer rates so different globally, and what do we do about it? Fertil Steril 2020;114:680–9. 10.1016/j.fertnstert.2020.09.003.

[15] Yu L, Lyu Y, Zhang F. Correlation between multiple embryo transfers and the incidence of preterm birth and low birth weight: a network meta-analysis. Arch Gynecol Obstet 2025;312:1049–61. 10.1007/s00404-025-08136-x.

[16] Eskew AM, Jungheim ES. A history of developments to improve in vitro fertilization. Mo Med 2017;114:156–9.

[17] Kanaka V, Proikakis S, Drakakis P, et al. Implementing a preimplantation proteomic approach to advance assisted reproduction technologies in the framework of predictive, preventive, and personalized medicine. EPMA J 2022;13:237–60. 10.1007/s13167-022-00282-5.

[18] Benkhalifa M, Madkour A, Louanjli N, et al. From global proteome profiling to single targeted molecules of follicular fluid and oocyte: contribution to embryo development and IVF outcome. Expert Rev Proteomics 2015;12:407–23. 10.1586/14789450.2015.1056782.

[19] Petricoin III EF, Ardekani AM, Hitt BA, et al. Use of proteomic patterns in serum to identify ovarian cancer. The Lancet 2;359:572.

[20] Davis JB, Andes S, Espina V. Reverse phase protein arrays. Methods Mol Biol Clifton NJ 2021;2237:103–22. 10.1007/978-1-0716-1064-0_9.

[21] Petricoin EF, Espina V, Araujo RP, et al. Phosphoprotein pathway mapping: Akt/mammalian target of rapamycin activation is negatively associated with childhood rhabdomyosarcoma survival. Cancer Res 2007;67:3431–40. 10.1158/0008-5472.CAN-06-1344.

[22] Gallagher RI, Wulfkuhle J, Wolf DM, et al. Protein signaling and drug target activation signatures to guide therapy prioritization: Therapeutic resistance and sensitivity in the I-SPY 2 Trial. Cell Rep Med 2023;4:101312. 10.1016/j.xcrm.2023.101312.

[23] Davuluri G, Espina V, Petricoin EF, et al. Activated VEGF receptor shed into the vitreous in eyes with wet AMD: a new class of biomarkers in the vitreous with potential for predicting the treatment timing and monitoring response. Arch Ophthalmol 2009;127:613–21. 10.1001/archophthalmol.2009.88.

[24] Butts CD, Bloom MS, McGough A, et al. Toxic elements in follicular fluid adversely influence the likelihood of pregnancy and live birth in women undergoing IVF. Hum Reprod Open 2021;2021:hoab023. 10.1093/hropen/hoab023.

[25] Levay PF, Huyser C, Fourie FLR, et al. The detection of blood contamination in human follicular fluid. J Assist Reprod Genet 1997;14:212–7.

[26] Fujimoto VY, Browne RW, Bloom MS, et al. Pathogenesis, developmental consequences, and clinical correlations of human embryo fragmentation. Fertil Steril 2011;95:1197–204. 10.1016/j.fertnstert.2010.11.033.

[27] Alikani M, Cohen J, Tomkin G, et al. Human embryo fragmentation in vitro and its implications for pregnancy and implantation. Fertil Steril 1999;71:836–42.

[28] Lewin A, Schenker JG, Safran A, et al. Embryo growth rate in vitro as an indicator of embryo quality in IVF cycles. J Assist Reprod Genet 1994;11:500–3.

[29] Holte J, Berglund L, Milton K, et al. Construction of an evidence-based integrated morphology cleavage embryo score for implantation potential of embryos scored and transferred on day 2 after oocyte retrieval. Hum Reprod 2007;22:548–57. 10.1093/humrep/del403.

[30] Hardarson T, Hanson C, Sjogren A, et al. Human embryos with unevenly sized blastomeres have lower pregnancy and implantation rates: indications for aneuploidy and multinucleation. Hum Reprod 2001;16:313–8. 10.1093/humrep/16.2.313.

[31] Gardner DK, Schoolcraft WB. Culture and transfer of human blastocysts. Curr Opin Obstet Gynecol 1999;11:307–11.

[32] Espina V, Mueller C. Solid pin protein array printing platforms. Adv Exp Med Biol 2019;1188:61–75. 10.1007/978-981-32-9755-5_4.

[33] Davidson B, Espina V, Steinberg SM, et al. Proteomic analysis of malignant ovarian cancer effusions as a tool for biologic and prognostic profiling. Clin Cancer Res Off J Am Assoc Cancer Res 2006;12:791–9. 10.1158/1078-0432.CCR-05-2516.

[34] Paria BC, Das SK, Dey SK. The preimplantation mouse embryo is a target for cannabinoid ligand-receptor signaling. Proc Natl Acad Sci 1995;92:9460–4. 10.1073/pnas.92.21.9460.

[35] Paria BC, Song H, Wang X, et al. Dysregulated cannabinoid signaling disrupts uterine receptivity for embryo implantation. J Biol Chem 2001;276:20523–8. 10.1074/jbc.M100679200.

[36] Walker OS, Holloway AC, Raha S. The role of the endocannabinoid system in female reproductive tissues. J Ovarian Res 2019;12:3. 10.1186/s13048-018-0478-9.

[37] Habara O, Logan CY, Kanai-Azuma M, et al. WNT signaling in pre-granulosa cells is required for ovarian folliculogenesis and female fertility. Development 2021;148:dev198846. 10.1242/dev.198846.

[38] Abedini A, Zamberlam G, Lapointe E, et al. WNT5a is required for normal ovarian follicle development and antagonizes gonadotropin responsiveness in granulosa cells by suppressing canonical WNT signaling. FASEB J 2016;30:1534–47. 10.1096/fj.15-280313.

[39] Vilotić A, Nacka-Aleksić M, Pirković A, et al. IL-6 and IL-8: an overview of their roles in healthy and pathological pregnancies. Int J Mol Sci 2022;23. 10.3390/ijms232314574.

[40] Liu T, Zhang L, Joo D, et al. NF-κB signaling in inflammation. Signal Transduct Target Ther 2017;2:17023. 10.1038/sigtrans.2017.23.

[41] Mauchart P, Vass RA, Nagy B, et al. Oxidative stress in assisted reproductive techniques, with a focus on an underestimated risk factor. Curr Issues Mol Biol 2023;45:1272–86. 10.3390/cimb45020083.

[42] Liu D, Xu Y. P53, oxidative stress, and aging. Antioxid Redox Signal 2011;15:1669–78. 10.1089/ars.2010.3644.

[43] Equils O, Kellogg C, McGregor J, et al. The role of the IL-1 system in pregnancy and the use of IL-1 system markers to identify women at risk for pregnancy complications. Biol Reprod 2020;103:684–94. 10.1093/biolre/ioaa102.

[44] Putri Susilo AF, Syam HH, Bayuaji H, et al. Free 25(OH)D3 levels in follicular ovarian fluid top-quality embryos are higher than non-top-quality embryos in the normoresponders group. Sci Rep 2024;14:29023. 10.1038/s41598-024-71769-6.

[45] Xu C, An X, Tang X, et al. Association between vitamin D level and clinical outcomes of assisted reproductive treatment: a systematic review and dose-response meta-analysis. Reprod Sci 2024. 10.1007/s43032-024-01578-9.

[46] Grzeczka A, Graczyk S, Skowronska A, et al. Relevance of vitamin D and its deficiency for the ovarian follicle and the oocyte: an update. Nutrients 2022;14. 10.3390/nu14183712.

[47] Krisher RL. The effect of oocyte quality on development. J Anim Sci 2004;82:E14–23. 10.2527/2004.8213_supplE14x.

[48] Sánchez F, Smitz J. Molecular control of oogenesis. Mol Genet Hum Reprod Fail 2012;1822:1896–912. 10.1016/j.bbadis.2012.05.013.

[49] Pan Y, Pan C, Zhang C. Unraveling the complexity of follicular fluid: insights into its composition, function, and clinical implications. J Ovarian Res 2024;17:237. 10.1186/s13048-024-01551-9.

[50] Dumesic DA, Meldrum DR, Katz-Jaffe MG, et al. Oocyte environment: follicular fluid and cumulus cells are critical for oocyte health. Fertil Steril 2015;103:303–16. 10.1016/j.fertnstert.2014.11.015.

[51] Alam MH, Miyano T. Interaction between growing oocytes and granulosa cells in vitro. Reprod Med Biol 2020;19:13–23. 10.1002/rmb2.12292.

[52] Gosden RG, Hunter RH, Telfer E, et al. Physiological factors underlying the formation of ovarian follicular fluid. J Reprod Fertil 1988;82:813–25. 10.1530/jrf.0.0820813.

[53] Shalgi R, Kraicer P, Rimon A, et al. Proteins of human follicular fluid: the blood-follicle barrier. Fertil Steril 1973;24:429–34. 10.1016/S0015-0282(16)39730-8.

[54] Wang C, Feng Y, Chen Y, et al. Proximity extension assay revealed novel inflammatory biomarkers for follicular development and ovarian function: a prospective controlled study combining serum and follicular fluid. Front Endocrinol 2025;16:1525392. 10.3389/fendo.2025.1525392.

[55] Hauser R, Mínguez-Alarcón L. Invited perspective: measuring biomarkers of exposure in target organs—what is needed to move forward? Environ Health Perspect 2023;131:061304. 10.1289/EHP12436.

[56] Kanaka V, Drakakis P, Loutradis D, et al. Proteomics in the study of female fertility: an update. Expert Rev Proteomics 2023;20:319–30. 10.1080/14789450.2023.2275683.

[57] Ouni E, Vertommen D, Amorim CA. The human ovary and future of fertility assessment in the post-genome era. Int J Mol Sci 2019;20. 10.3390/ijms20174209.

[58] Nishizuka S, Charboneau L, Young L, et al. Proteomic profiling of the NCI-60 cancer cell lines using new high-density reverse-phase lysate microarrays. Proc Natl Acad Sci 2003;100:14229–34. 10.1073/pnas.2331323100.

[59] Bianchi L, Gagliardi A, Landi C, et al. Protein pathways working in human follicular fluid: the future for tailored IVF? Expert Rev Mol Med 2016;18:e9. 10.1017/erm.2016.4.

[60] Zamah AM, Hassis ME, Albertolle ME, et al. Proteomic analysis of human follicular fluid from fertile women. Clin Proteomics 2015;12:5–5. 10.1186/s12014-015-9077-6.

[61] Shen X, Liu X, Zhu P, et al. Proteomic analysis of human follicular fluid associated with successful in vitro fertilization. Reprod Biol Endocrinol 2017;15:58. 10.1186/s12958-017-0277-y.

[62] Pla I, Sanchez A, Pors SE, et al. Proteome of fluid from human ovarian small antral follicles reveals insights in folliculogenesis and oocyte maturation. Hum Reprod 2021;36:756–70. 10.1093/humrep/deaa335.

[63] Przewocki J, Kossinski D, Lukaszuk A, et al. Follicular fluid proteomic analysis to identify predictive markers of normal embryonic development. Int J Mol Sci 2024;25:8431. 10.3390/ijms25158431.

[64] Galazis N, Olaleye O, Haoula Z, et al. Proteomic biomarkers for ovarian cancer risk in women with polycystic ovary syndrome: a systematic review and biomarker database integration. Fertil Steril 2012;98:1590-1601.e1. 10.1016/j.fertnstert.2012.08.002.

[65] Lu Y, Ling S, Hegde AM, et al. Using reverse-phase protein arrays as pharmacodynamic assays for functional proteomics, biomarker discovery, and drug development in cancer. Pharmacodyn Cancer Drug Dev 2016;43:476–83. 10.1053/j.seminoncol.2016.06.005.

